# DAVE: how to use explainable AI to interpret missense variants for genome diagnostics based on functional protein modeling

**DOI:** 10.1101/2025.11.25.25340947

**Authors:** Tim Niemeijer, René Mulder, Helga Westers, Jan D.H. Jongbloed, Bart Charbon, Birgit Sikkema-Raddatz, Lennart F. Johansson, Marielle E. van Gijn, Cleo C. van Diemen, Dennis Hendriksen, Kristin M. Abbott, W.T. Kars Maassen, Morris A. Swertz, K. Joeri van der Velde

## Abstract

Diagnostic yield in NGS genome diagnostics is constraint by the high fraction of variants of uncertain significance (VUS), in large part due to insufficient interpretability of missense variation. Existing pathogenicity predictors offer strong performance, but often produce an unexplainable score lacking mechanistic insight. Here, we present the Digital Approximation of Variant Effects (MOLGENIS DAVE), an explainable missense variant predictor built on 12 biophysically grounded features spanning stability, hydrophobicity, electrostatics, and molecular interactions. Trained on curated Dutch diagnostic data, DAVE reliably classifies and breaks down predictions into interpretable feature contributions. With a focus on explainability, this framework aims to alleviate the VUS burden, advances clinically actionable variant interpretation and enables mechanistic follow-up. All source code used to process and integrate the data, along with the data required to reproduce all annotations and analyses, is available at https://github.com/molgenis/dave.

## 1 Introduction

### 1.1 Missense is still a bottleneck in genome diagnostics

Despite decades of active research, the diagnostic yield of next-generation sequencing (NGS) in genome diagnostic analysis rarely exceeds 35%^1–6^. A leading cause is the large fraction of DNA variants classified as variants of uncertain significance (VUS)^7^. Particularly missense variants are notoriously difficult to interpret as a result of their subtle and context-dependent effects on protein function. According to established clinical guidelines^8^, determining whether a missense variant is pathogenic requires a combination of multiple strong lines of evidence and supporting data. However, these criteria are often only partially met, leading to uncertainty in classification^9,10^. Additional strong evidence from follow-up functional studies can resolve VUS by directly assessing the mechanistic consequence of individual variants. Yet, such studies are laborious, variant specific, and given the sheer volume of variants identified in NGS genome diagnostics, often impractical^11^.

### 1.2 Most predictors lack interpretable insights

To aid in prioritizing and classifying missense variants, a multitude of pathogenicity predictors have been developed^12^. These predictors employ a range of approaches, including evolutionary conservation^13^, allele frequency data^14^, ensemble learning methods^15^, and training labels derived from evolutionary ^16^ or clinical sources^17^. Other strategies focus on the impact of variants on protein domains^18^ or structural features^19^, while some emphasize visualization to aid interpretation^20^. Currently, there are over 400 such predictors^21^. And while the field has started to suffer from data leakage^22^, imbalance^23^, bias, and circularity^24,25^, few predictors have attempted to combine genomics with less biased physical modeling^26^.The few methods that do, lack explainability ^27–30^ or are highly disease specific^31,32^.

AlphaFold^33,34^ represents a major breakthrough in our ability to simulate the molecular effects of amino acid substitutions by dramatically increasing the number of high-confidence protein structure models across the human proteome^35^. An unbiased and high-performance missense pathogenicity predictor based on AlphaFold protein models along with evolutionary data and contextual features is AlphaMissense^36^. However, like most predictors, it produces only numerical predictions without explanations. This lack of interpretability limits our ability to investigate the underlying causal mechanisms, which is essential for translating computational predictions into clinically actionable insights and to guide functional laboratory experiments.

### 1.3 Using explainable AI to interpret pathogenicity predictions

We hypothesize that the gap between traditional predictive modeling and mechanistic evidence can be bridged by breaking down prediction output into comprehensible feature contributions that allow further functional interpretation. This requires selecting biologically relevant features for the predictor, derived from protein stability, hydrophobicity, electrostatics, and interaction with ligands, DNA, RNA, and other proteins, combined with insight into the model’s decision making provided by SHapley Additive exPlanations (SHAP)^37^ that quantify the contribution of each feature to individual predictions. The result can guide targeted digital or experimental follow-up tests to confirm or refute pathogenicity and help prioritize missense variants of interest. In this manuscript, we describe how we tested this hypothesis by developing Digital Approximation of Variant Effects, part of the MOLGENIS software family (MOLGENIS DAVE) to predict pathogenicity on carefully selected functionally relevant features, trained on pathogenic and benign variants from accredited Dutch genome diagnostic laboratories. We applied this model to the VUS of this dataset and compared the predictions with a more recent release of the same dataset and classifications in ClinVar^38^. Lastly, we break down the selected predictions and highlight three examples in which explainability assists in their mechanistic evaluation.

## 2 Results

### 2.1 The DAVE prediction model

Functional features were computed for a selection of missense variants from the April 2024 release of fully de-identified and publicly accessible variant classifications provided by all Dutch genome diagnostic laboratories through the Datashare working group of the Dutch Clinical Genetics Laboratory Society (VKGL)^39^. From these, we selected 12 biophysically grounded features from five complementary sources: P2Rank^40^, FoldX 5^41^, GLM-Score^42^, Peptides R package^43^ and GeoNet^44^, as summarized in table 1. The DAVE model was trained on the feature differences (i.e., delta) between wild-type and protein structures containing an amino acid substitution caused by benign (LB/B) and pathogenic (LP/P) missense variants split into a train and test dataset. We determined an ROC AUC of 86% on the test set and a probability of 0.286 as the most effective binary classification threshold to identify pathogenic variants.

**Table 1:** The 12 features used for training the DAVE model. References to tools and structures are provided within the table for additional information.

| Name | Unit | Source | Note |
| --- | --- | --- | --- |
| Folding energy $\Delta G$ | kJ/mol | FoldX5<sup>41</sup> | Sum of 15 Gibbs free energy change terms |
| Net electric charge at pH 7 | pKa | R-Peptides<sup>43</sup> | Henderson-Hasselbalch, Lehninger scale |
| Hydrophobic moment | $\mu H$ | R-Peptides<sup>43</sup> | Amphipathicity at $\phi=100^\circ$ in 11AA fractions |
| Hydrophobicity | GRAVY | R-Peptides<sup>43</sup> | Non-polar affinity on Kyte-Doolittle scale |
| Isoelectric point | pH | R-Peptides<sup>43</sup> | Computed on the EMBOSS pK scale |
| Ligand binding no. pockets | pockets | P2Rank<sup>40</sup> | Total no. of potential binding pockets |
| Ligand binding top pocket | SAS point | P2Rank<sup>40</sup> | Nr. 1 pocket solvent accessible surface |
| DNA binding affinity | pKd | GLM-Score<sup>42</sup> | Based on binding RCSB PDB entry 1BNA<sup>45</sup> |
| RNA binding affinity | pKd | GLM-Score<sup>42</sup> | Based on binding RCSB PDB entry 4JRD<sup>46</sup> |
| DNA binding site | residue | GeoNet<sup>44</sup> | Uses geometry, residue-DNA binding patt. |
| RNA binding site | residue | GeoNet<sup>44</sup> | Uses geometry, residue-RNA binding patt. |
| Protein binding site | residue | GeoNet<sup>44</sup> | Uses geometry, residue-prot binding patt. |

### 2.2 Retrospective classification of VUS

The trained DAVE model was applied on VUS missense variants from the VKGL April 2024 data release. Using the binary classification threshold, 3,801 variants were characterized as pathogenic and 7,420 as benign. To compare DAVE’s classification performance with real-world reclassification of VUS, we used a more recent release of the VKGL dataset and variant classifications from ClinVar. In July 2025, compared to April 2024, only 12 VUS had been reclassified as likely pathogenic/pathogenic or likely benign/benign on the VKGL data-sharing platform. These variants are listed in table 2. Of these, nine DAVE predictions were in concordance with the re-classification, of which four true positives (i.e., correctly identified as pathogenic) and five true negatives (i.e., correctly identified as benign). Two were false positives (i.e., incorrectly predicted as pathogenic), and one was false negative (i.e., incorrectly predicted as benign). Additionally, cross-referencing of the VKGL VUS with ClinVar revealed that 478 variants were classified as benign/likely benign, while 176 were considered likely pathogenic/pathogenic. Figure 1 illustrates the distribution of DAVE pathogenicity probabilities and the corresponding reclassification labels from VKGL and ClinVar.

**Figure 1:**
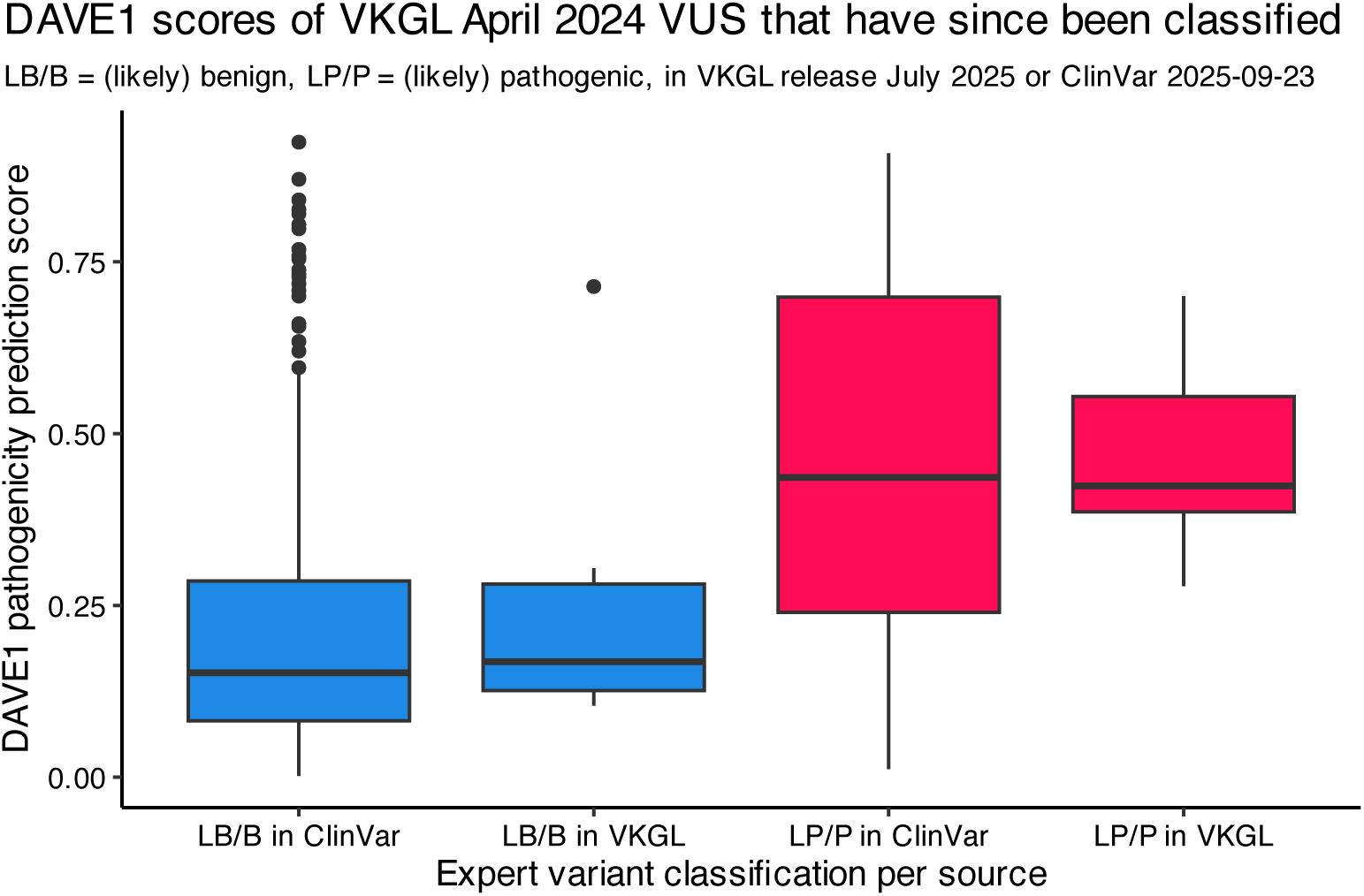
Box plots of DAVE pathogenicity probabilities for 654 ClinVar and 12 VKGL retrospectively classified variants. These variants were VUS in VKGL release April 2024 and have since been reclassified, categorized by classification label (LB/LP) and classification source.

**Table 2:**
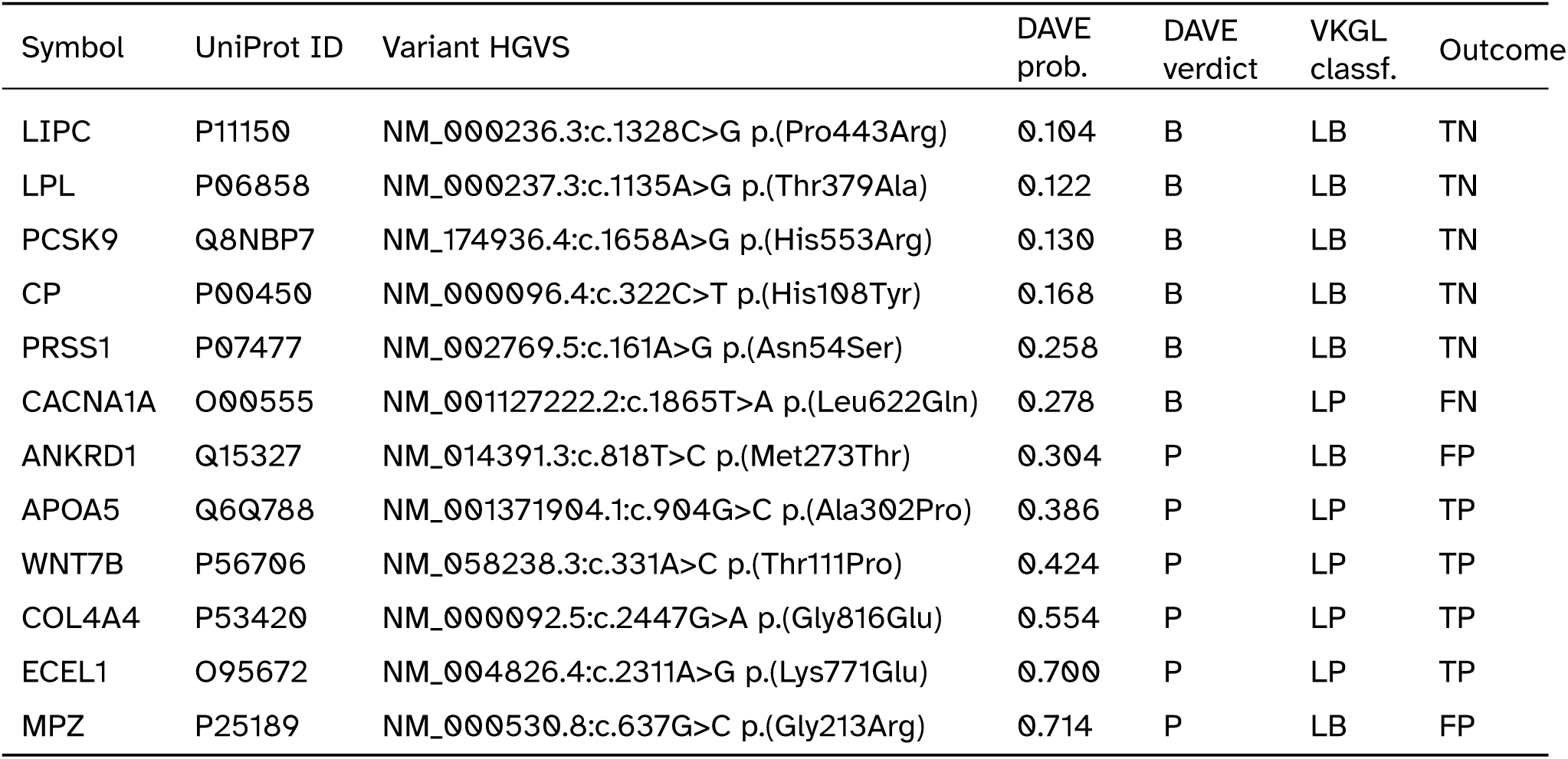
The 12 VKGL VUS missense variants that have received an updated classification in a more recent release of the VKGL dataset. These variants were VUS in VKGL release April 2024 and have since been reclassified by the VKGL in release July 2025. DAVE pathogenicity probabilities and verdict, based on a threshold of 0.286, are shown. Outcomes: TN = True Negative, FN = False Negative, TP = True Positive, FP = False Positive.

| Symbol | UniProt ID | Variant HGVS | DAVE prob. | DAVE verdict | VKGL classf. | Outcome |
| --- | --- | --- | --- | --- | --- | --- |
| LIPC | P11150 | NM_000236.3:c.1328C>G p.(Pro443Arg) | 0.104 | B | LB | TN |
| LPL | P06858 | NM_000237.3:c.1135A>G p.(Thr379Ala) | 0.122 | B | LB | TN |
| PCSK9 | Q8NBP7 | NM_174936.4:c.1658A>G p.(His553Arg) | 0.130 | B | LB | TN |
| CP | P00450 | NM_000096.4:c.322C>T p.(His108Tyr) | 0.168 | B | LB | TN |
| PRSS1 | P07477 | NM_002769.5:c.161A>G p.(Asn54Ser) | 0.258 | B | LB | TN |
| CACNA1A | O00555 | NM_001127222.2:c.1865T>A p.(Leu622Gln) | 0.278 | B | LP | FN |
| ANKRD1 | Q15327 | NM_014391.3:c.818T>C p.(Met273Thr) | 0.304 | P | LB | FP |
| APOA5 | Q6Q788 | NM_001371904.1:c.904G>C p.(Ala302Pro) | 0.386 | P | LP | TP |
| WNT7B | P56706 | NM_058238.3:c.331A>C p.(Thr111Pro) | 0.424 | P | LP | TP |
| COL4A4 | P53420 | NM_000092.5:c.2447G>A p.(Gly816Glu) | 0.554 | P | LP | TP |
| ECEL1 | O95672 | NM_004826.4:c.2311A>G p.(Lys771Glu) | 0.700 | P | LP | TP |
| MPZ | P25189 | NM_000530.8:c.637G>C p.(Gly213Arg) | 0.714 | P | LB | FP |

### 2.3 Functional interpretation

To demonstrate how DAVE can guide interpretation of functional consequences, we performed a more detailed analysis of three VUS that were reclassified in the July 2025 VKGL data-sharing release compared to the April 2024 release or had a different classification in ClinVar. These analyses are enabled by DAVE’s ability to break down the final pathogenicity probability into separate feature contributions as SHAP values^37^, capturing interactions unique to each prediction. SHAP values of contributing features, including their delta and unit, are shown in a cumulative plot per variant. ChimeraX^47^ was used to visualize the predicted effects on protein structures.

#### 2.3.1 *WNT7B* T111P

The variant in *WNT7B* NM_058238.3:c.331A>C causes the amino acid substitution Thr111Pro in the Wnt-7b protein. It was selected for having the highest absolute folding energy change of the variants with an updated classification in the VKGL dataset in table 2). It is predicted to have a substantial increase in ΔG folding energy, see figure 2. A higher ΔG causes protein folding to occur less spontaneous, affecting correct folding and stability. Molecular visualization shows a change in the hydrogen bond integral in the secondary structures and their connecting elements (figure 3). The impact on protein structural integrity can be a good mechanistic explanation of the effect of this missense variant.

**Figure 2:**
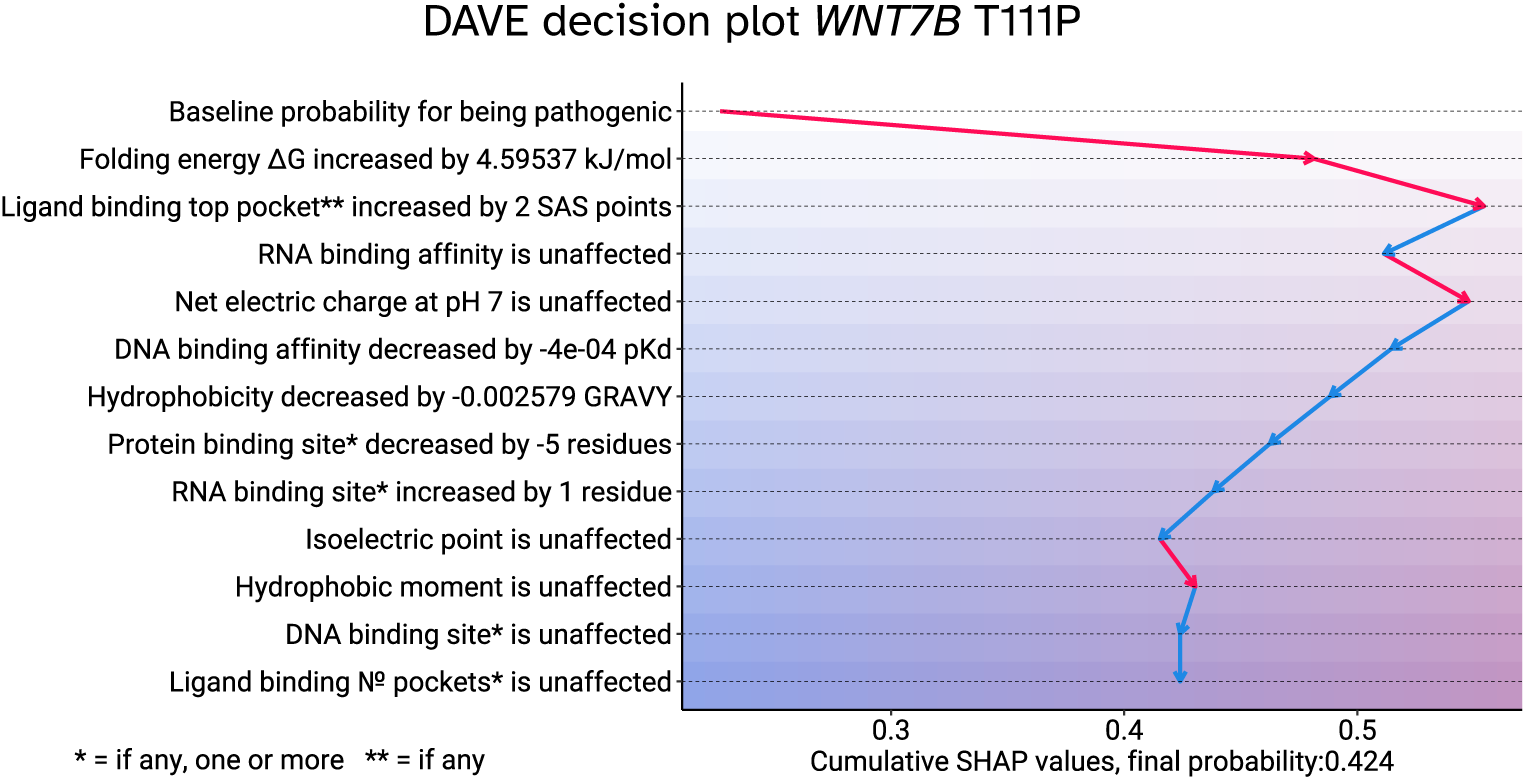
DAVE decision plot illustrating the predicted impact of the T111P amino acid substitution resulting from a variant in *WNT7B*. Features are on the y-axis, starting with a baseline pathogenicity probability, followed by contributing features ranked by their impact. Cumulative SHAP probability contributions are shown with arrows on the x-axis, ending on the final pathogenicity probability at the bottom.

**Figure 3:**
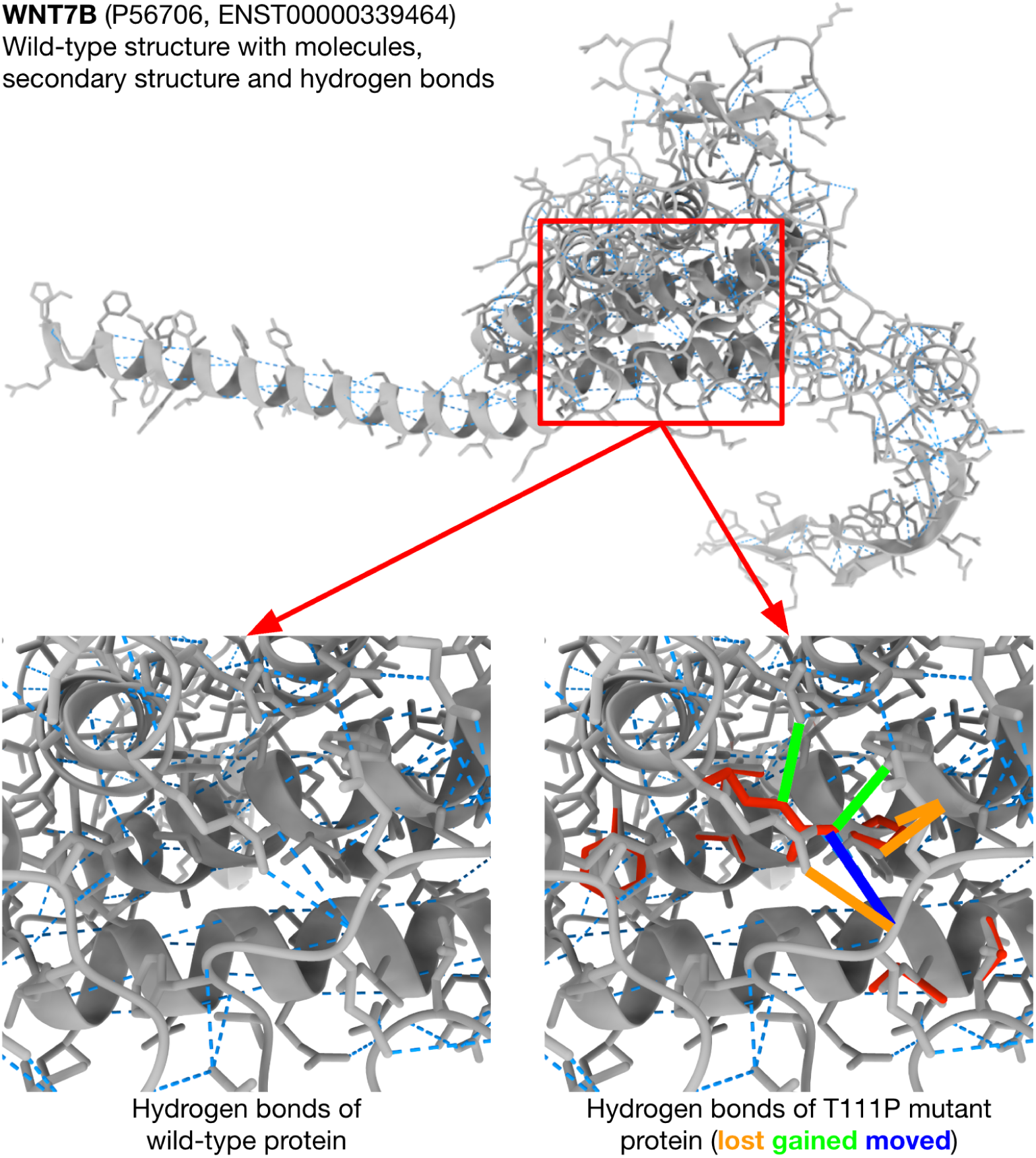
Structural comparison of wild-type protein encoded by *WNT7B* to the variant protein with a T111P substitution. The complete protein wild-type structure is shown on top with, indicated with the red box are the affected region of the variant. Cropped images of the indicated region are shown at the bottom left and at the bottom right. The bottom left showing the wild-type structure. At the bottom right a cropped image of wild-type is shown with the T111P structure overlay in red. Hydrogen bonds are indicated with blue dotted lines with changes highlighted with solid lines.

#### 2.3.2 *SLCO2A1* G554R

The SLCO2A1 variant NM_005630.2:c.1660G>A, causes the amino acid substitution Gly554Arg in the solute carrier organic anion transporter family member 1A2 protein. It was selected for having the largest absolute change in solvent-accessible surface (SAS) points within its top-ranking ligand-binding pocket. Although currently still classified as a VUS in the VKGL dataset, it is reported as pathogenic in ClinVar^48^. DAVE predicted the variant to be pathogenic, see figure 4). The predicted differences in the electrostatic surface potential of the ligand-binding pocket in the simulated structure of the G554R versus wild-type protein, are shown in figure 5. Moreover, recent functional characterization of variants in the transmembrane domain of *SLCO2A1* replacing glycine with residues with larger side chains underscored this, as it revealed a loss of helical dynamics, accompanied by potential disruption of the helical packing^49^.

**Figure 4:**
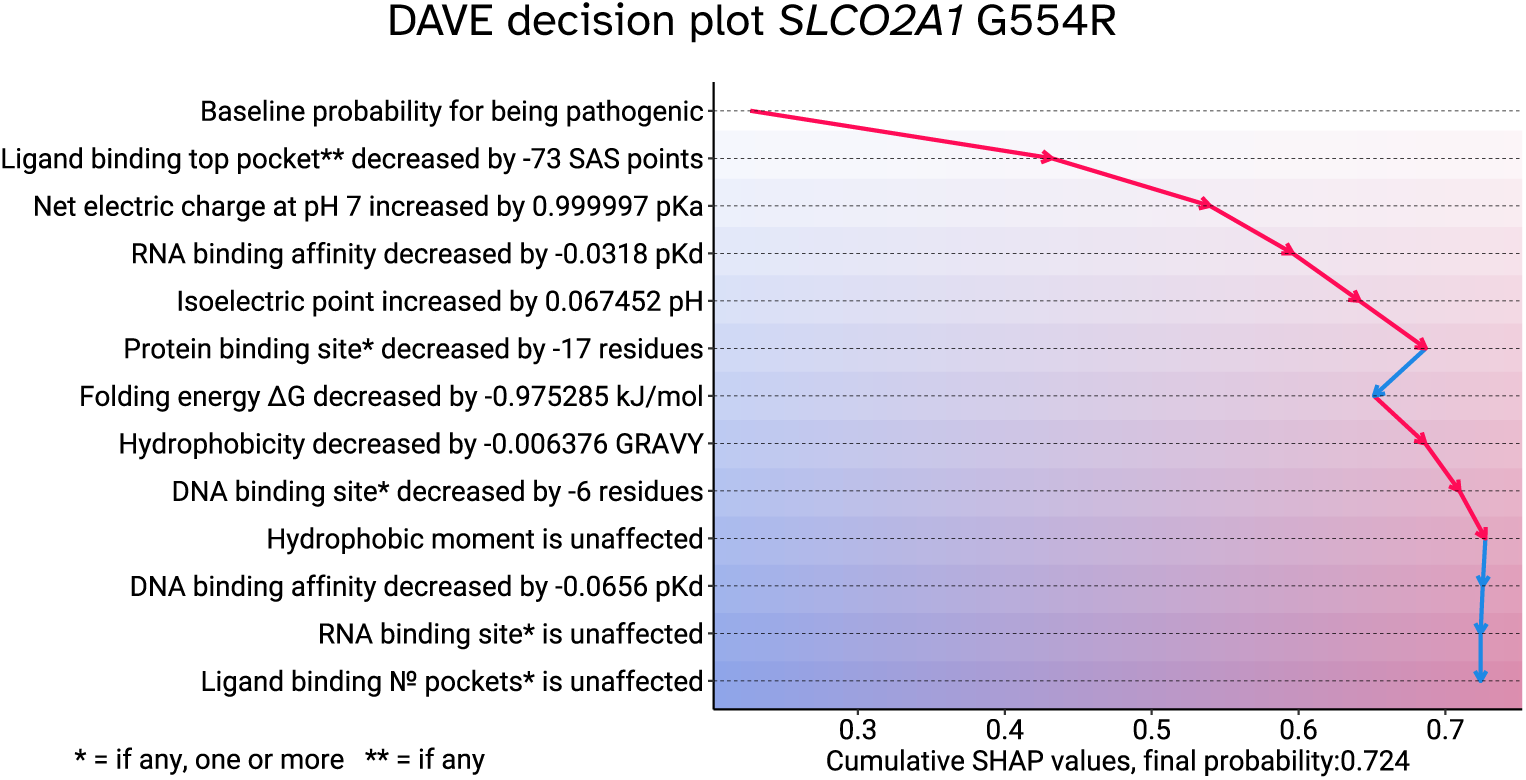
DAVE decision plot illustrating the predicted impact of the G554R amino acid substitution resulting from a variant in *SLCO2A1*. Features are on the y-axis, starting with a baseline pathogenicity probability, followed by contributing features ranked by their impact. Cumulative SHAP probability contributions are shown with arrows on the x-axis, ending on the final pathogenicity probability at the bottom.

**Figure 5:**
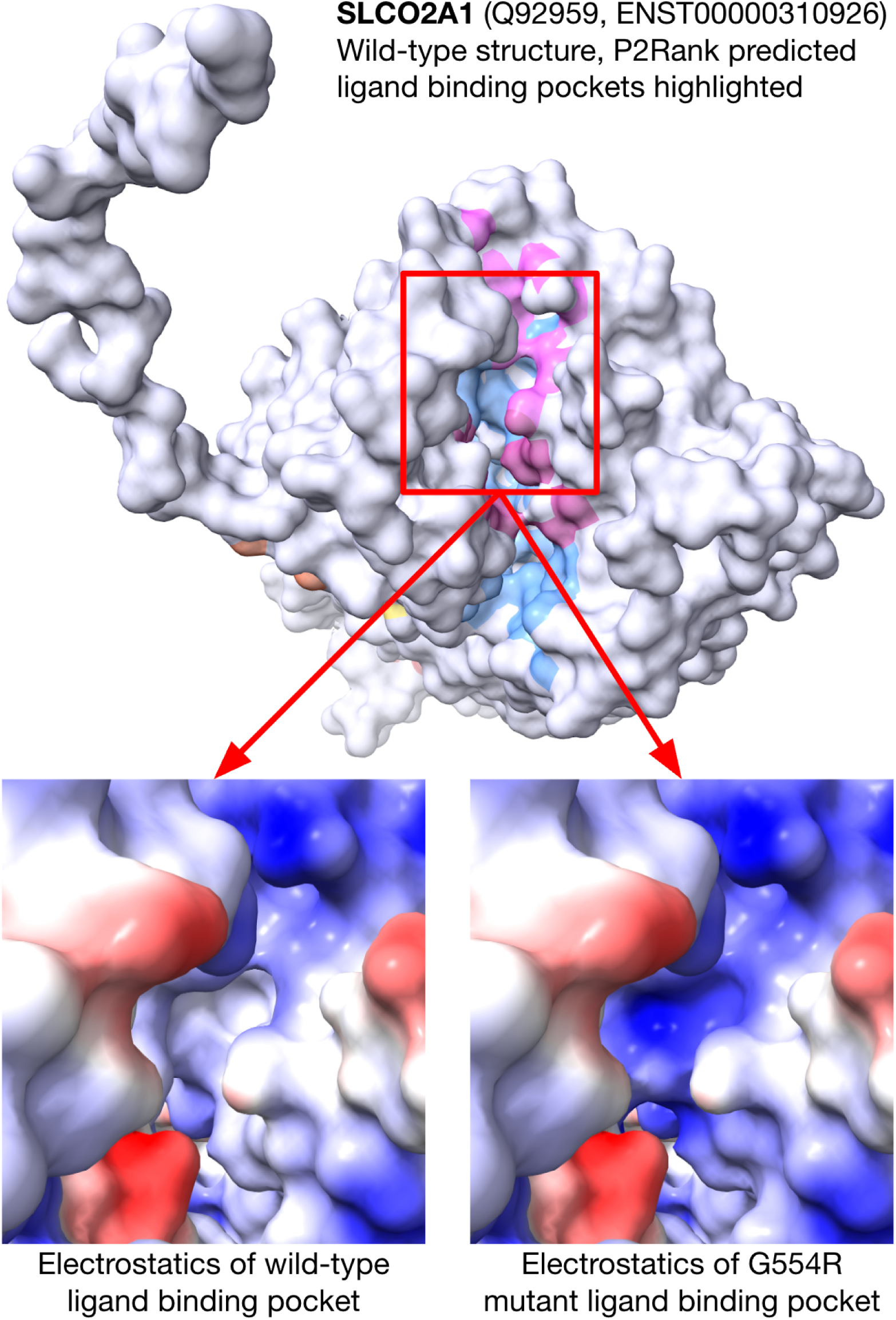
Structural comparison of wild-type protein encoded by *SLCO2A1* to the protein with the G554R substitution. Top: wild-type protein is displayed in white, with pink, light-blue, orange and yellow accents given to the predicted ligand binding pockets. The region of interest is indicated with the red box. Bottom: the Coulombic electrostatic surface potential is shown for region of interest, where red indicates negative potential and blue is positive potential.

#### 2.3.3 *NKX2-5* L153P

The variant *NKX2-5* NM_004387.3:c.458T>C, causes the amino acid change, Leu153Pro in the homeobox Nkx-2.5 protein. It was selected for having largest absolute change in protein binding site residues. This variant is currently classified as VUS in the VKGL dataset while reported as pathogenic in ClinVar. DAVE predicts that this variant is pathogenic, see figure 6. The protein encoded by *NKX2-5* is a transcription factor, and the substitution L153P, lies in an important homeodomain (AA 138-197) responsible for the binding of DNA as well as other transcription factors^50^. Interestingly, despite the location of the variant, the most contributing feature to the predicted pathogenicity of this variant is not the change in binding site residues, but the change in folding energy ΔG. The predicted structural change of the L153P variant protein compared to the wild-type is visualized in figure 7.

**Figure 6:**
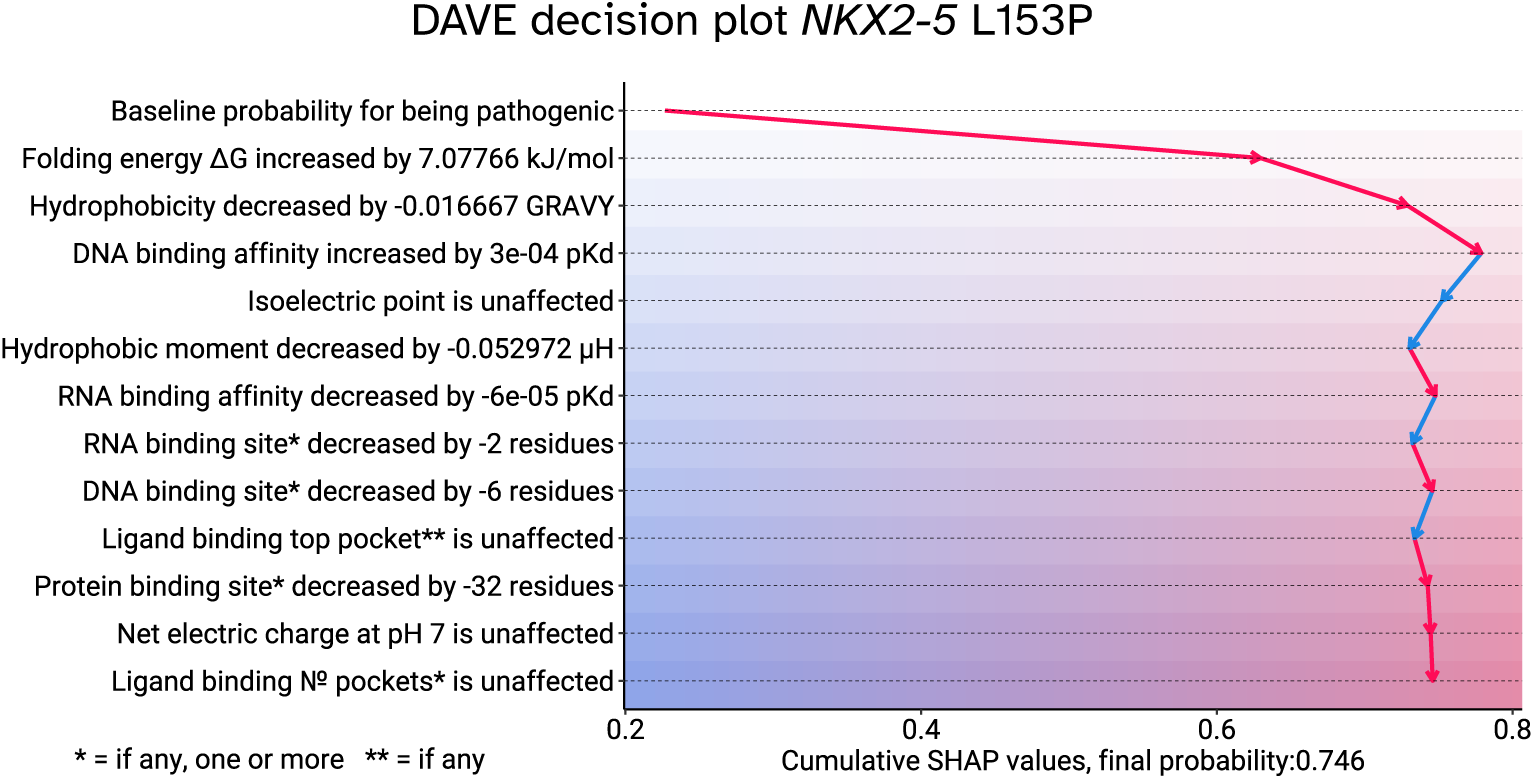
DAVE decision plot illustrating the predicted impact of the L153 amino acid substitution resulting from a variant in *NKX2-5*. Features are on the y-axis, starting with a baseline pathogenicity probability, followed by contributing features ranked by their impact. Cumulative SHAP probability contributions are shown with arrows on the x-axis, ending on the final pathogenicity probability at the bottom.

**Figure 7:**
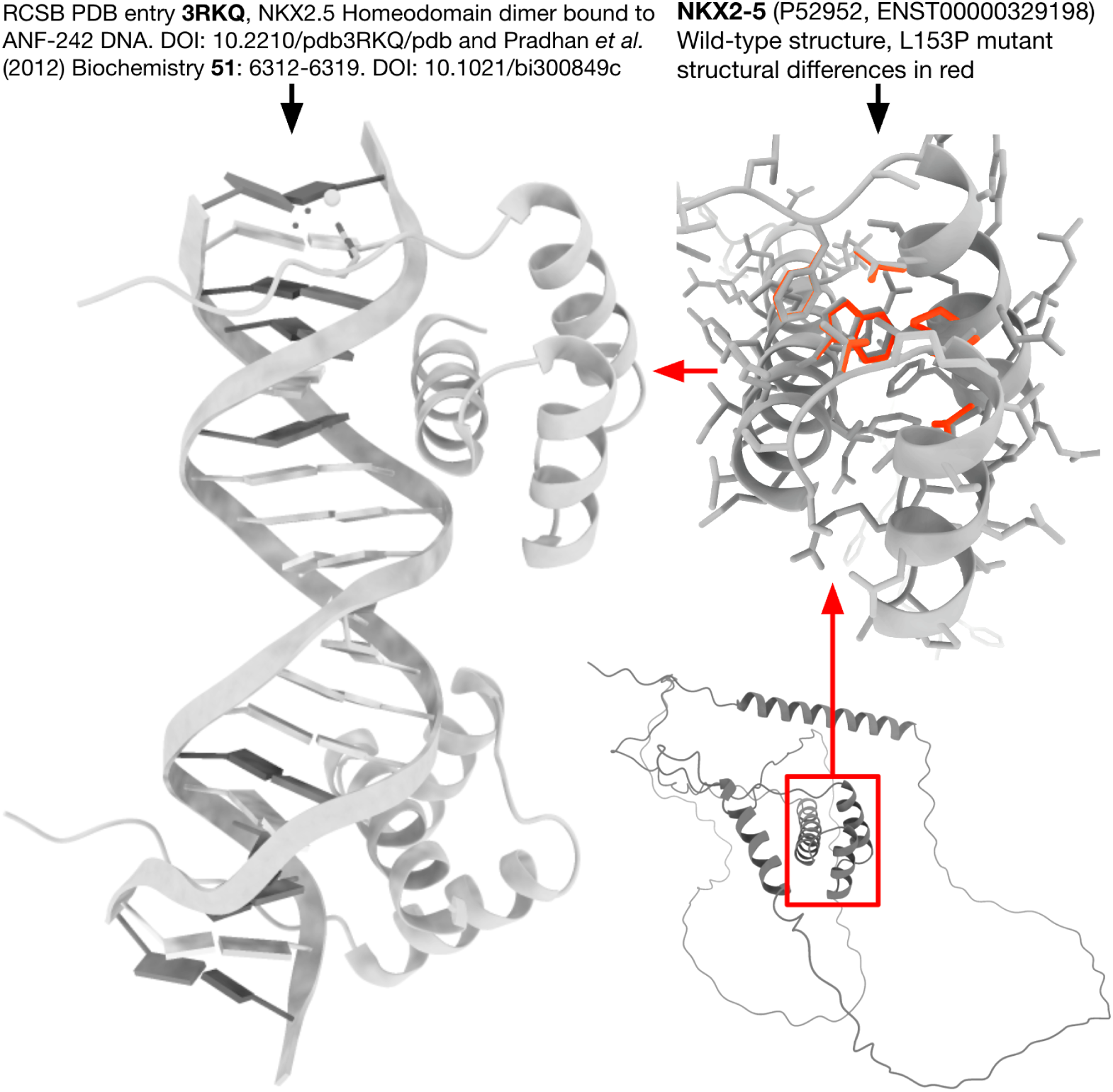
Structural comparison of the protein with the L153P substitution to the wild-type protein encoded by the *NKX2-5* gene. Left: Homeodomain (HD) interacting with ANF-242 DNA sequence. Right bottom: Highlighted in the red box, the location of the HD in the protein encoded by *NKX2-5*. Right top: HD where L153P structure changes are shown in red.

## 3 Discussion

### 3.1 DAVE successfully prioritizes physical explainability

In this study, we developed MOLGENIS DAVE, a supervised learning model that predicts pathogenicity of missense variants and provides interpretable insights via contributions of functionally relevant features based on protein modeling. Unlike methods such as AlphaMissense^7^ and REVEL^15^, DAVE prioritizes physical explainability over predictive power, providing unique and complementary evidence for variant classification and guidance for targeted experimental validation of VUS either in dry or wet laboratory.

### 3.2 Evaluation of features in the DAVE model

The selected features of DAVE represent multiple disease-causing mechanisms^51^ and thus provide a robust starting point for interpretation. However, no single model can fully capture the complexity and unique biology of all proteins. Manual follow-up will remain essential to accurately determine the molecular consequences of prioritized variants. By integrating more diverse and context-dependent features, future versions of DAVE could better capture the nuanced mechanisms underlying variant effects, ultimately improving both accuracy and interpretability across a broader range of proteins and variant types. For instance, variants that have a gain-of-function or dominant-negative consequence have different, often milder, molecular mechanisms compared to loss-of-function variants^52,53^. Other examples of such effects include allostery, post-translational modifications (PTMs), toxicity, structural flexibility, folding rate, kinetic effects, aggregation propensity, membrane association, immunogenicity, and co-operativity.

### 3.3 Limitations in current protein structure

Some AlphaFold structures are predicted with low confidence, particularly for disordered regions or poorly conserved domains^54,55^. In these regions, predictive accuracy can be significantly lower, especially if models rely heavily on structural data^56^. Another limitation of current models is that they represent the protein without their biological context, and that models larger than 2700 residues (<2% of all models) are chunked into smaller fragments due to computational limitations, making them unsuitable for meaningful predictions. To increase applicability across diverse protein types and missense effect types, structure confidence metrics and more biological context should be integrated to refine functional predictions. We expect that future versions of AlphaFold and similar efforts will produce unfragmented models for all proteins.

### 3.4 Overcoming technical barriers for broad applicability

While DAVE could certainly be configured to run as part of DNA interpretation pipelines, currently it is not offered an easy to install standalone product. The reasons are the integration with external software, one of which used under academic personal license, as well as a prohibitive computational burden, averaging around 20 minutes per variant on commodity hardware. This hinders its applicability in clinical genetics workflows where rapid turnaround time is essential. For future versions, we will prioritize improving computational efficiency and ensuring permissive licensing of underlying methods.

### 3.5 Conclusion and future perspectives

In summary, DAVE combines predictive accuracy with meaningful information on how its predictions are made. By complementing numerical predictions with mechanistic insights, we demonstrate the potential to transform in silico variant effect predictions into interpretable molecularly grounded explanations. The insights provide a practical route to generate testable hypotheses for the resolution of VUS in functional follow-up studies, thereby bridging the gap between prediction, explanation, and clinical utility. Looking ahead, we envision a framework that integrates DAVE with complementary evidence such as population frequency, conservation, inheritance, and phenotypes as well as specialized tools that cover effects on transcription and translation to systematically classify missense variants and reduce VUS. The importance of using an ensemble of dedicated tools to cover specific effects is exemplified by our false negative prediction for *CACNA1A* Leu622Gln, see table 2. This variant was classified in diagnostics as likely pathogenic due to a predicted impact on splicing, which explains why DAVE did not predict a significant pathogenic effect at the protein level. Integrating DAVE into pipelines such as MOLGENIS VIP^57^ would enable the inclusion of these complementary tools within a scalable and modular infrastructure for variant interpretation. Such an integrative approach enables large scale reanalysis efforts, to prioritize variants for experimental validation, and ultimately accelerate the route from variant discovery to informed clinical decision making.

## 4 Methods

### 4.1 Datasets

The main resource is the fully de-identified and publicly accessible DNA variant classifications dataset collected in April 2024 by the Datashare working group of the nine genome diagnostic labs in the Netherlands, organized in the VKGL (Vereniging Klinisch Genetische Laboratoriumdiagnostiek)^39^. We downloaded the GRCh38 liftover version from the MOLGENIS VIP^57^ resources. If variants within the same gene with the same protein change had differences in classification, they were marked as conflicting. The July 2025 version of the VKGL release, was collected similarly to the April 2024 release. For protein structures, we used pdb’s from the Al-phaFold v4 release. AlphaFold structures are represented by UniProt^58^ and mappings to HGNC gene symbols are provided. The GRCh38 VCF file for ClinVar^38^ release 2025-09-23 was used as-is by matching on chromosome, position, reference and alternative allele.

### 4.2 Data processing

The VKGL variant dataset was annotated with Ensembl VEP^59^ version 112. Based on protein localization data from the Protein Atlas^60^ we randomly selected 1007 intracellular, 1028 membrane, and 692 secreted proteins for which we also had genomic variants in the dataset. This selection of proteins was used to subset the VKGL variants to balance the data on localization. Additionally, we annotated whether chaperones were involved in the folding of these proteins, for this we used a dataset of 194 manually curated chaperones^61^ and expanded these genes with UniProt/SwissProt IDs using Ensembl BioMart ^62^. These known chaperones were connected to their interaction protein partners using BioGRID^63^.

We applied the following five protein analysis tools on the selected variants, P2Rank^40^, FoldX 5^41^, GLM-Score^42^, Peptides R package^43^, and GeoNet^44^. Complete data processing was successful for 23,417 variants, of which the classifications, localizations, and chaperoned folding characteristics are shown in table 3.

**Table 3:**
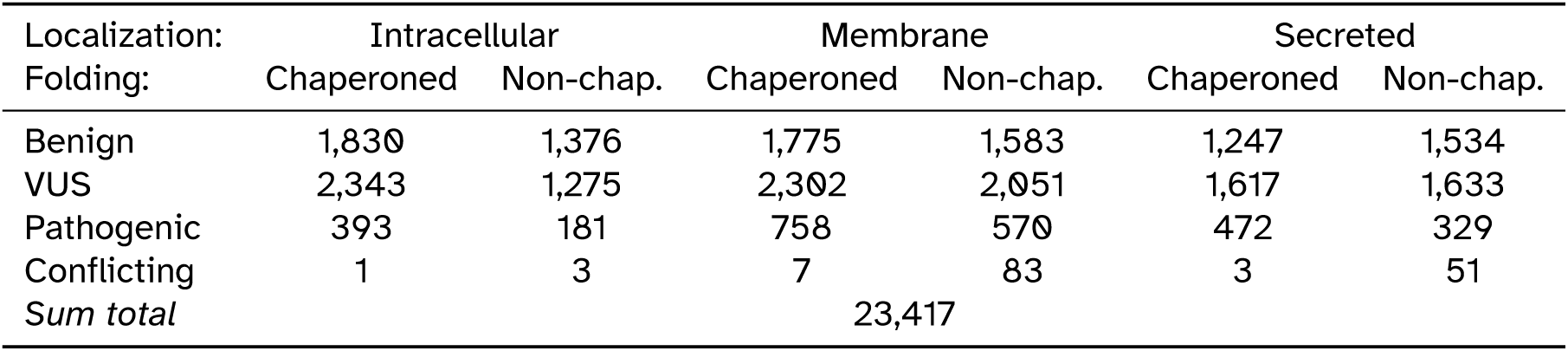
D**a**ta **used to train, test and apply the DAVE model.** Each number represents a set of DNA variants randomly sampled from the VKGL release of April 2024, though equalized across protein localization and folding pathway. Benign includes ’likely benign’ and ’benign’, Pathogenic includes ’likely pathogenic’ and ’pathogenic’. VUS are variants of uncertain significance.

| Localization: | Intracellular |  | Membrane |  | Secreted |  |
| --- | --- | --- | --- | --- | --- | --- |
| Folding: | Chaperoned | Non-chap. | Chaperoned | Non-chap. | Chaperoned | Non-chap. |
| Benign | 1,830 | 1,376 | 1,775 | 1,583 | 1,247 | 1,534 |
| VUS | 2,343 | 1,275 | 2,302 | 2,051 | 1,617 | 1,633 |
| Pathogenic | 393 | 181 | 758 | 570 | 472 | 329 |
| Conflicting | 1 | 3 | 7 | 83 | 3 | 51 |
| *Sum total* | 23,417 |  |  |  |  |  |

### 4.3 Feature engineering

To obtain the most informative and independently contributing set of features, the set was minimized by removing correlations and selecting the most informative and explainable features. Among the remaining features, pairwise correlations were minimal, with the exception of correlation between ’Net electric charge at pH 7’ and ’Isoelectric point’. Despite this, both features were deemed valuable to retain due to their individual relevance. The R function ’cor’ from the stats (r-core) package was used to calculate the correlation of features as the Pearson correlation coefficient (PCC), shown in figure 8.

**Figure 8:**
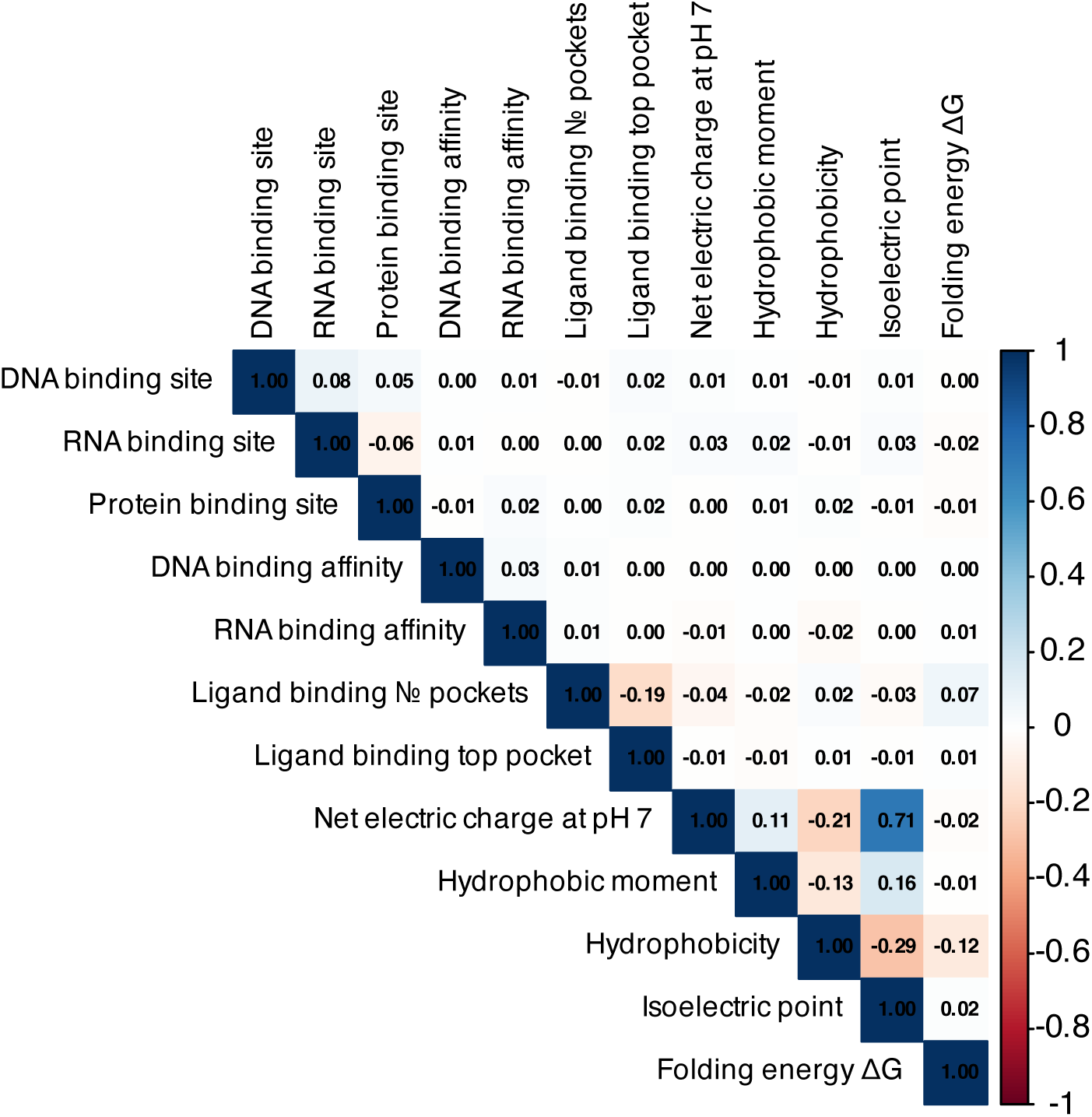
**Feature correlation in the DAVE training data**. Correlation was calculated as the Pearson correlation coefficient.

### 4.4 Model training

A subset of the full annotated dataset, consisting of 12,048 variants classified as likely pathogenic (LP) or likely benign (LB), was split into training and testing sets using an 80/20 ratio. The DAVE model was implemented using the randomForest package in R (version 4.7-1.1) ^64^ using the standard Random Forest algorithm with default parameters. To determine the contribution of each selected feature to individual predictions, SHAP values were calculated using the R package fastshap^65^(version 0.1.1) with 10-fold Monte Carlo sampling. It is important to note that SHAP values do not correlate with feature values, but instead capture feature contributions based on the interactions among all features uniquely for this prediction.

### 4.5 Optimal classification threshold

For binary classification, the VKGL April 2024 ’test’ set was used to determine the optimal threshold with the R package cutpointr ^66^(version 1.1.2). From this, a threshold of 0.286 was established as the cutoff point using Youden’s J statistic^67^ expressed as:

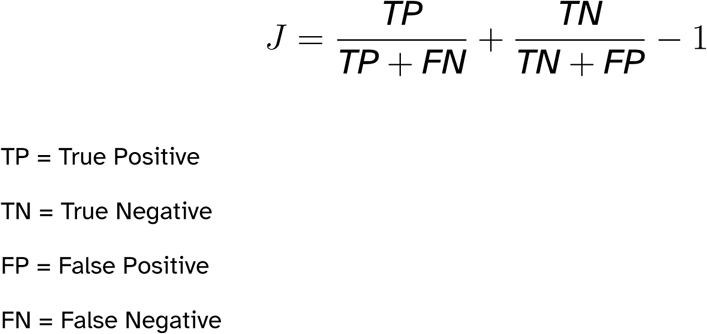

### 4.6 Retrospective comparison

To evaluate the utility of DAVE in the reclassification of the 11,221 variants of uncertain significance (VUS), we compared variant classifications between the April 2024 and July 2025 VKGL datasets. Additionally, we compared variants classified as VUS in the VKGL that had a B/LB or LP/P classification in ClinVar (September 23, 2025, with at least a one star review status and no conflicting classifications) with the predictions of DAVE. For variant specific structural comparison, we substituted wild-type amino acid with the variant in the AlphaFold^33^ structure .pdbs using FoldX5^41^ with the BuildModel and RepairPDB command. These structures were then visualized with ChimeraX^47^ and compared with wild-type structures.

## Declarations

### Ethics approval and consent to participate

Not applicable.

### Consent for publication

Not applicable.

### Availability of data and materials

The April 2024 VKGL public datashare variants were downloaded from https://download.m olgeniscloud.org/downloads/vip/resources/GRCh38/vkgl_consensus_20240401.tsv the original GRCh37 version is available via https://vkgl.molgeniscloud.org. Protein localization data were downloaded from: https://www.proteinatlas.org/about/download Chaperone interaction datasets were downloaded from the supplement of Shemesh et al, from https://static-content.springer.com/esm/art%3A10.1038%2Fs41467-021-22369-9/MediaObjects/41467_2021_22369_MOESM3_ESM.xlsx and BIOGRID ORGANISM 4.4.234 tab3 from https://downloads.thebiogrid.org/BioGRID/Release-Archive/BIOGRID-4.4.234/ AlphaFold (version 4) structures were downloaded from: https://ftp.ebi.ac.uk/pub/da tabases/alphafold/v4/UP000005640_9606_HUMAN_v4.tar All training/testing data and predictions of the reinterpreted VUS are available at https://github.com/molgenis/DAVE. The DAVE results for VKGL VUS data are also available through an interactive dashboard at https://dave.molgeniscloud.org/.

### Availability of source code

All code is available as free and open source software under the GNU Lesser General Public License (LGPL) v3.0 at https://github.com/molgenis/dave. P2Rank^40^ is available at https://github.com/rdk/p2rank. FoldX 5^41^ is available under academic license at https://foldxsuite.crg.eu/. GLM-Score^42^ is available at https://github.com/Klab-Bioinfo-Tools/GLM-Score. Peptides R package^43^ is available at https://cran.r-project.org/web/packages/ Peptides. GeoNet^44^, the version we used is available at https://github.com/joerivandervelde/GeoNet.

### Author contributions

T.N., R.M., H.W., J.D.H.J. and K.J.V. conceived the project. T.N. performed the functional interpretation. K.J.V. performed the data processing. T.N. and K.J.V. wrote the manuscript with critical input and revisions from R.M., H.W., J.D.H.J., B.C., B.S.R., L.F.J., M.E.G., C.C.D., D.H., K.M.A., W.T.K.M., and M.A.S. All authors have reviewed and approved the manuscript.

## Data Availability

All data produced are available online at https://github.com/molgenis/dave

https://download.molgeniscloud.org/downloads/vip/resources/GRCh38/vkgl_consensus_20240401.tsv

https://vkgl.molgeniscloud.org

https://www.proteinatlas.org/about/download

https://static-content.springer.com/esm/art%3A10.1038%2Fs41467-021-22369-9/MediaObjects/41467_2021_22369_MOESM3_ESM.xlsx

https://downloads.thebiogrid.org/BioGRID/Release-Archive/BIOGRID-4.4.234/

https://ftp.ebi.ac.uk/pub/databases/alphafold/v4/UP000005640_9606_HUMAN_v4.tar

## Acknowledgements

We would like to thank our colleagues at Genomics Coordination Center for engineering support and access to the UMCG high performance compute cluster. We are grateful to our colleagues from all Dutch genome diagnostic laboratories for their continuing efforts to share variant classifications through the VKGL Datashare working group. We would also like to thank our previous and current heads of department Prof. Nine Knoers and Prof. Anke-Hilse Maitland-van der Zee as well as the UMCG Board of Directors, in particular Prof. Stephanie Klein Nagelvoort Schuit and Prof. Wiro Niessen for their encouragement and support to develop and use new AI methods at the University Medical Center Groningen. Lastly, we would like to express our gratitude to the exquAIro AI bootcamp organization and board members Prof. Gerard Koppelman, Dr. Martin Smit, Ilya Petoukhov, Dr. Marnix Bügel, as well as Geerte Koster of REWIRE and Dr. Kai Yu Ma of UMCG for their training, coaching and valuable input.

